# Newborn Life Support Course: Does it make me more confident when resuscitating a newborn?

**DOI:** 10.1101/2020.08.04.20114637

**Authors:** Laurent Renesme, Maria Merched, Olivier Tandonnet, Julien Naud

## Abstract

**Objective:** To describe the effectiveness of the Neonatal Life Support (NLS) course in terms of attendees’ knowledge, perceived self-efficacy, and clinical applicability.

**Methods:** We conducted an electronic survey of NLS course attendees (NLS+ group). The survey had six themes: i) demographic characteristics; ii) NLS clinical applicability; iii) attendee’s perceived proficiency at neonatal resuscitation; iv) attendee’s perceived experience of fluency, security, and quality of care during neonatal resuscitation; v) knowledge (multiple-choice questions); and vi) perceived personal and professional impact of the NLS course. A control group (NLS−) was recruited via our regional perinatal network. The survey data were analysed anonymously. Multiple linear regression analysis examined the following: NLS course, job tenure, maternity level, and profession.

**Results:** The survey completion rate was 62% (200/323) for the NLS+ group. Among participants, 84% had participated in neonatal resuscitation since their course. The scores for positive perceived experience for neonatal resuscitation (fluency, security, and quality of care delivered) were higher in the NLS+ group than the NLS− group (*p* < 0.006). After adjustment, the independent factors associated with a higher positive perceived experience were the NLS course, work in tertiary level maternity ward, and job tenure > 5 years. The multiple-choice questions score (n = 10) was 8.2 ± 1.3 (NLS+) *vs*. 6.7 ± 1.5 (NLS−) (*p* < 0.0001). NLS course, medical degree, and work in a tertiary level maternity ward were independently associated with higher knowledge scores.

**Conclusion:** The NLS course was associated with better knowledge of, and a positive perceived experience regarding, neonatal resuscitation.

## Introduction

Adaptation to the extra-uterine environment is a major physiological transition. Some newborns, especially preterm infants, require intervention at birth, most often as non-invasive respiratory support ^1^. Management of these newborns is paramount, as poor adaptation at birth, despite its low incidence, is associated with high morbidity and mortality ^2–4^

The European Resuscitation Council (ERC) developed the Newborn Life Support (NLS) course based on international recommendations from the International Liaison Committee on Resuscitation (ILCOR) and ERC ^5^. This course is widely taught in Europe. However, only a few studies have evaluated its impact, focusing mainly on the attendees’ performance at technical procedures ^6–8^.

Therefore, we assessed the effectiveness of the NLS course in terms of the attendees’ knowledge, perceived self-efficacy, and clinical applicability.

## Methods

### Study population

We enrolled two groups from among perinatal healthcare professionals from the same geographic region (Nouvelle Aquitaine, France). NLS course (NLS+ group) attendees were recruited via our local NLS course attendee email list (years 2014–2018). A control group (NLS−) were recruited via our regional perinatal network mailing list.

### Survey design

We developed an anonymous self-assessment questionnaire based on Artino *et al*. ^9,10^. The items assessing the participants’ perceived experience were answers using a 5-level Likert scale. Three NLS experts reviewed the initial survey to ensure its validity and proper understanding of the items. The survey was administered using Google Form; reminder emails were send 7 and 14 days after dissemination of the online survey.

The NLS+ group survey included 48 items divided into six parts. The first part included the participants’ demographic characteristics. The second asked the participants to assess the applicability of the NLS course in their practice and if they had participated in neonatal resuscitation since the NLS course. For each positive response, the participants had to describe their perceived experience. The third asked the participants about their perceived proficiency with 10 items for neonatal resuscitation at birth (*e.g*. positive-pressure ventilation, chest compressions, etc.). In the fourth, the participants evaluated their perceived fluency, security, and quality of care during neonatal resuscitation. The fifth assessed theoretical knowledge with 10 multiple-choice questions (MCQs) from the NLS manual. Part six assessed the professional and personal impact of the NLS course: its applicability to professional life and improvement of professional qualifications and personal development. The NLS− controls were given the same survey minus parts 2 and 6.

### Data analysis

Quantitative variables are presented as the mean ± standard deviation. For items scored with Likert scales, the results were transformed into numerical values ranging from 1 (worst appreciation) to 5 (best appreciation). Qualitative variables are presented as percentages.

Univariate analysis was performed using the Mann–Whitney *U-test* (quantitative variables) or χ^2^ test (qualitative variables). Multiple linear regression analysis was used to analyse the following exposure variables: NLS course (NLS+ *vs*. NLS−), job tenure (≤ 5 vs. > 5 years), maternity level (tertiary vs. primary or secondary), and profession (physician vs. paramedic and midwives).

The statistical analyses were carried out using R ^11^.

## Results

### Study populations

For the NLS+ group, 200 surveys were completed by 323 NLS course attendees from 2014 to 2018 (completion rate 62%). The control NLS− group completed 65 surveys. There were no missing data in the surveys. Table 1 presents the demographic characteristics of the NLS+ and NLS− groups. Forty-six (23%) participants from the NLS+ group declared having attended another neonatal resuscitation training, compared to 47 (72%) in the NLS-group (p <.0001).

**Table 1.** Demographic characteristics.

|  |  | **NLS+ group**<br>**n=200** | **NLS- group**<br>**n=65** | **p value** |
| --- | --- | --- | --- | --- |
| **Age (year)**<br>(mean ± standard deviation) | | 35 ± 9.4 | 42 ± 11.2 | <.0001 |
| **Male**<br>n (%) |  | 25 (12) | 8 (12) | 0.97 |
| **Profession**<br>n (%) | Paramedic | 25 (12) | 6 (9) | 0.48 |
|  | Midwife | 100 (50) | 47 (72) | 0.002 |
|  | Physician | 75 (38) | 12 (19) | 0.0045 |
| **Maternity level**<br>n (%) | Primary | 43 (22) | 22 (34) | 0.04 |
|  | Secondary | 44 (22) | 27 (41) | 0.002 |
|  | Tertiary | 88 (44) | 7 (11) | <.0001 |
|  | Structure with no maternity | 25 (12) | 9 (14) | 0.78 |
| **Job tenure (year)**<br>(mean ± standard deviation) | | 9.7 ± 8.5 | 16.8 ± 10.7 | <.0001 |

Univariate analysis was performed using the Mann–Whitney *U*-test (quantitative variables) or $\chi^2$ test (qualitative variables).

### NLS applicability in professional work (NLS+ group)

Since their NLS course, 84% of the respondents had participated in neonatal resuscitation in the delivery room, in 48% of the cases as the leader. Participation in emergency procedures included upper airway management (82%), positive-pressure ventilation (63%), umbilical venous catheter insertion (31%), chest compressions (27%), and adrenaline administration (18%).

The perceived experience score for fluency (/5) when performing technical procedures was as follows: leadership 3.9 ± 0.7, upper airway management 4.3 ± 0.7, positive-pressure ventilation 4.2 ± 0.8, chest compressions 4.1 ± 0.9, umbilical venous catheter insertion 4.6 ± 0.6, and adrenaline administration 4.4 ± 0.8.

Participants declared that the NLS course has good applicability in their professional life (4.1 ± 1) and believed that NLS had improved their professional qualifications (4.2 ± 0.8) and personal development (4.3 ± 0.8).

### Theoretical knowledge

The MCQ score (/10) was 8.2 ± 1.3 for the NLS+ group *vs*. 6.7 ± 1.5 for the NLS− controls (*p* < 0.0001).

### Perceived proficiencies and experience

Participants’ perceived proficiencies for 10 items for neonatal resuscitation at birth are presented in table 2.

**Table 2.** Perceived proficiencies.

|  | **NLS+ group**<br>**n=200** | **NLS- group**<br>**n=65** | **p value** |
| --- | --- | --- | --- |
| Initial assessment and support of transition. | 4.4 ± 0.7 | 4.5 ± 0.8 | 0.2 |
| Prevent hypothermia. | 4.6 ± 0.6 | 4.5 ± 0.8 | 0.2 |
| Leadership. | 3.7 ± 0.9 | 3.3 ± 1 | 0.03 |
| Upper airway management. | 4.5 ± 0.6 | 4.2 ± 0.9 | 0.03 |
| Positive pressure ventilation. | 4.3 ± 0.7 | 4.0 ± 1.1 | 0.02 |
| Positive pressure ventilation efficacy evaluation.<br>Strategies to correct inadequate ventilation. | 4.0 ± 0.8 | 3.6 ± 1.1 | 0.03 |
| Chest compressions. | 4.0 ± 0.8 | 3.7 ± 1.1 | 0.3 |
| Umbilical venous catheter insertion. | 3.4 ± 1.3 | 2.1 ± 1.4 | <0.0001 |
| Adrenaline administration. | 3.7 ± 1.1 | 3.6 ± 1.2 | 0.5 |
| Tracheal intubation. | 2.7 ± 1.3 | 2.7 ± 1.3 | 0.9 |

mean ± standard deviation.
Univariate analysis (Mann–Whitney *U*-test).

The respective average scores for perceived fluency, security, and quality of care during neonatal resuscitation (/5) for the NLS+ and NLS− groups were 3.6 ± 0.8 vs. 3.1 ± 1.1 (*p* = 0.0002), 3.6 ± 0.9 vs. 3.1 ± 1 (*p* < 0.0001), and 3.7 ± 0.9 vs. 3.3 ± 1 (*p* = 0.0006). Table 3 shows the results of the multiple linear regression analysis for perceived experience (fluency, security and quality of care during neonatal resuscitation) and theoretical knowledge (MCQ score). After adjustment, NLS course, work in tertiary level maternity ward, and job tenure > 5 years were independent factors associated with a more positive perceived experience. NLS course, medical profession, and work in a tertiary level maternity ward were independent factors associated with better theoretical knowledge.

**Table 3.**
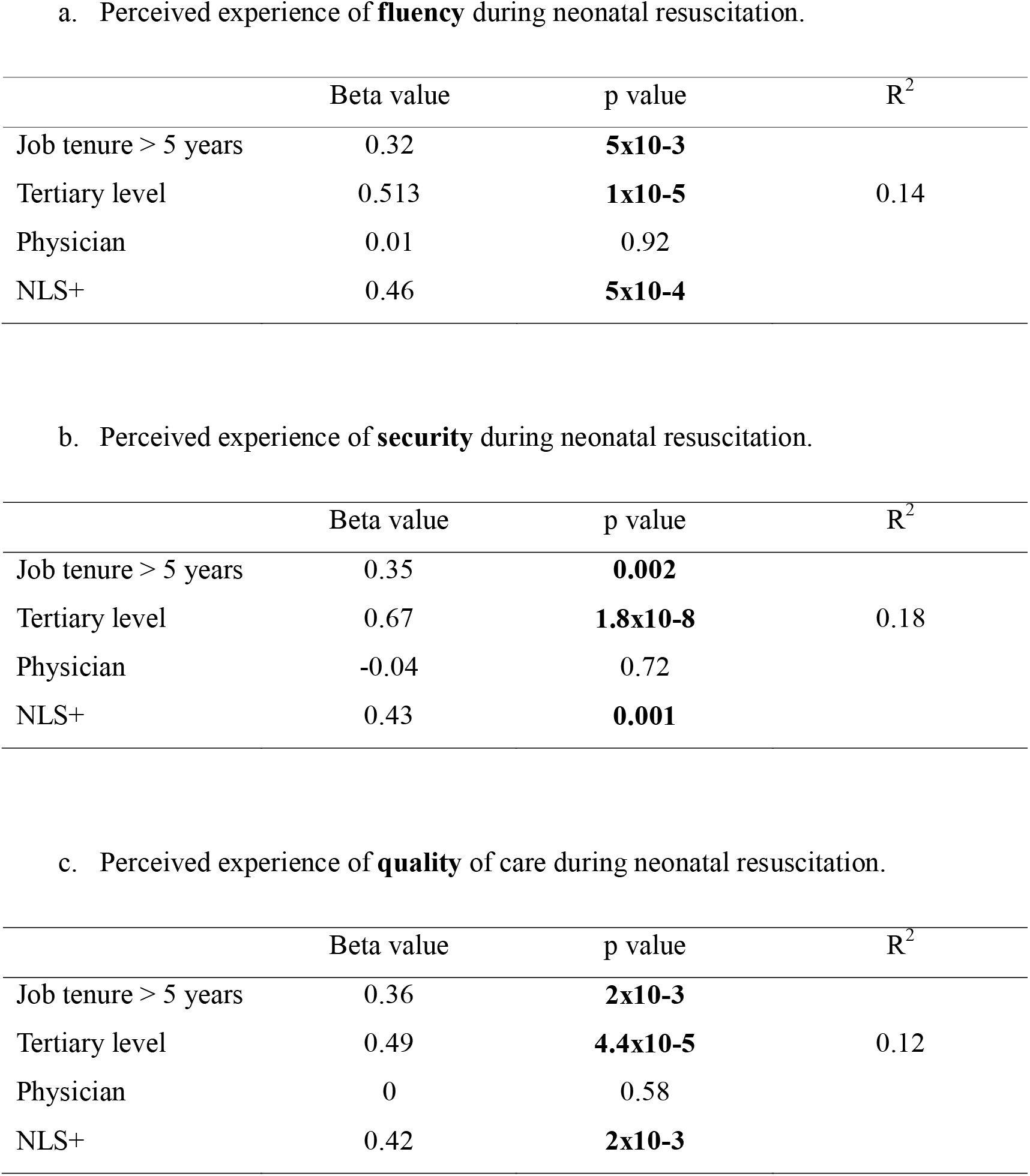
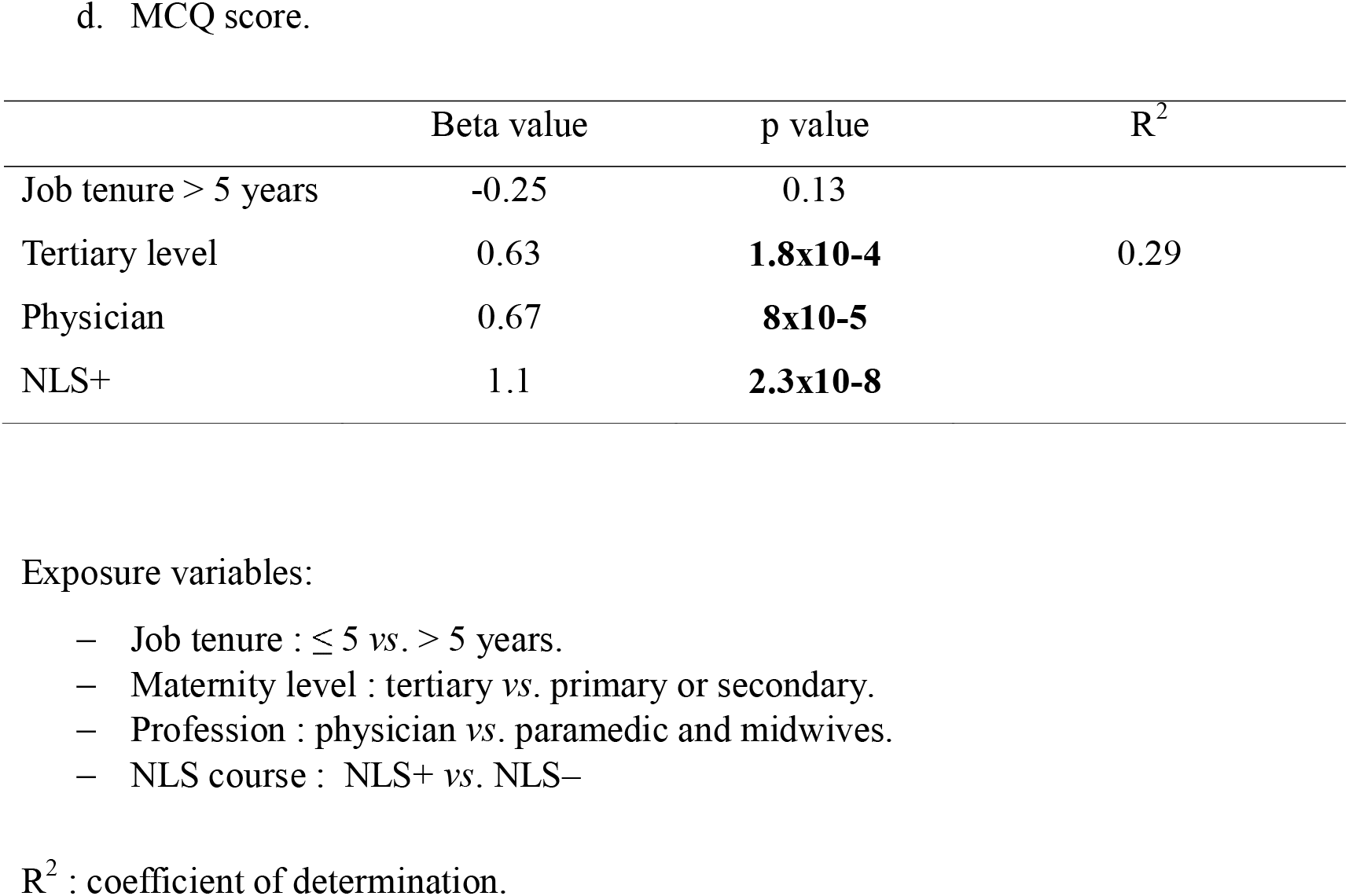
Multiple linear regression analysis.

## Discussion

In our study, the majority of NLS course attendees had participated in one or more neonatal resuscitations in the delivery room since their initial course (84%). The NLS course was associated with a positive perceived experience of fluency (> 4/5) for all technical procedures. Participation in the NLS course was an independent predictor of a positive perceived experience in terms of fluency, safety, quality of care delivered, and better theoretical knowledge.

Three studies have evaluated the NLS course and its impact, assessing technical skills on a manikin ^6^, upper airway management and theoretical knowledge ^7^, and positive-pressure ventilation and theoretical knowledge ^8^. The NLS course was associated with better knowledge and improved ventilation technique. However, these skills deteriorate over time. Evaluation of a similar course (the Neonatal Resuscitation Program) also found discrepancies between technical skills and knowledge, with deterioration in technical skills shortly after training and better retention of knowledge ^12^.

The originality of our study was that it focused on the perceived experience of NLS attendees, *i.e*. self-efficacy, a cognitive process indicating people’s confidence in their ability to effect a given behaviour ^13^. This performance-based confidence can be distinguished from both theoretical knowledge and technical skills necessary to perform resuscitation. Self-efficacy seems to be important for the success of resuscitation. Healthcare professionals who are knowledgeable and qualified in technical resuscitation procedures may not apply them successfully if they do not have a sufficiently strong belief in their own abilities ^14^ The data in the literature are discordant regarding self-efficacy and skills self-assessment ^7,14,15^ However, Olson *et al*. showed that training in neonatal resuscitation procedures improves the feeling of self-efficacy ^16^. The higher this feeling of self-efficacy was, the more caregivers tended to use positive-pressure ventilation at birth. This emphasised the importance of a positive perceived experience in terms of “self-efficacy” to be able to feel comfortable and undertake neonatal resuscitation.

The main limitation of our study is the small number of participants in the control group, as compared to the NLS+ group. However, this selection bias was limited by adjustment (multiple linear regression analysis).

## Conclusion

Participation in an NLS course was associated with a positive perceived experience in terms of fluency, safety, and quality of care delivered in the event of neonatal resuscitation in the delivery room and with a better theoretical knowledge. Based on our results, the NLS course seems to strengthen the feeling of “self-efficacy” in healthcare professionals, which is critical to performing neonatal resuscitation.

The English in this document has been checked by at least two professional editors, both native speakers of English. For a certificate, please see: http://www.textcheck.com/certificate/r5hKX1

## Data Availability

Data from this study are not available.

## Ethics

The Ethics Committee of the University Hospital of Bordeaux emitted a favourable recommendation for this study (Reference GP-CE2020-24).

## Acknowledgments

The authors want to acknowledge Pr J. Harambat for his advice and help with the statistical analysis.

